# Differences in the serum metabolomic profile of progressive alcohol-related liver disease in comparison to non-progressive alcohol-related liver disease: a cross sectional metabolomics study

**DOI:** 10.1101/2024.12.10.24318756

**Authors:** Eemeli Puhakka, Hany Ahmed, Retu Haikonen, Sophie Leclercq, Kati Hanhineva, Luca Maccioni, Camille Amadieu, Marko Lehtonen, Ville Männistö, Jaana Rysä, Peter Stärkel, Olli Kärkkäinen

## Abstract

Alcohol-related liver disease (ALD) is a major cause of mortality and disability adjusted life years. It is not fully understood why a small proportion of patients develop progressive forms of ALD (e.g. fibrosis, cirrhosis). Differences in the metabolic processes could be behind the individual progression of ALD. Our aim was to examine differences in serum metabolome between patients with non-progressive ALD and patients with an early form of progressive ALD.

The study had three study groups: progressive ALD (alcohol-related steatohepatitis or early-stage fibrosis, n=50), non-progressive ALD (simple steatosis, n=50) and healthy controls (n=32). Both ALD groups took part in a voluntary alcohol rehabilitation program. A non-targeted metabolomics analysis and targeted analysis of short chain fatty acids was done to the serum samples taken on the day of admission.

We found 111 significantly (p<0.0005) altered identified metabolites between the study groups. Our main finding was that levels of glycine conjugated bile acids, glutamic acid, 7-methylguanine and several phosphatidylcholines were elevated in the progressive ALD group in comparison to both the non-progressive ALD group and the controls. Glycine conjugated bile acid, glutamic acid and 7-methylguanine also positively correlated with increased levels of aspartate aminotransferase, alanine aminotransferase, gamma-glutamyl transferase, cell death biomarker M65, and liver stiffness.

Our results indicate that the enterohepatic cycle of glycine conjugated bile acids as well as lipid and energy metabolism are altered in early forms of progressive ALD. These metabolic processes could be a target for preventing progression of ALD.

## Introduction

Alcohol use is one of the leading causes of disability adjusted life years and mortality worldwide (Bryazka et al. 2022). Alcohol-related liver disease (ALD) is one of the most prevalent liver diseases and the leading cause of liver cirrhosis worldwide (Devarbhavi et al. 2023). Most (80-90%) chronic heavy drinkers develop some degree of ALD (Ohashi et al. 2018, Mathurin and Bataller 2015). ALD related mortality and disease burden has been on the rise in both Europe and the United States in recent years (Åberg et al. 2024, Moon et al. 2020).

ALD encompasses a spectrum of diseases, ranging from benign isolated steatosis to the more advanced stages steatohepatitis, cirrhosis and eventually liver cancer. The early stage (i.e. steatosis) is mostly asymptomatic and completely reversible with abstinence from alcohol (Åberg et al. 2020). Although most ALD cases present primarily as steatosis, only 20-40 % of those progress to steatohepatitis and 8-20% further develop cirrhosis (Ohashi et al. 2018, Mathurin and Bataller 2015) and eventually hepatocellular carcinoma (HCC) (Ganne-Carrié and Nahon 2019). Late stages of ALD (i.e., cirrhosis, and HCC) carry a poor prognosis (Ginès et al. 2021) with HCC accounting for around 80% of all primary liver cancer patients (Toh et al. 2023).

The early detection of ALD and its progression are hindered by the lack of specificity and sensitivity in traditional liver injury and alcohol biomarkers, such as aspartate aminotransferase (AST), alanine aminotransferase (ALT) and gamma-glutamyltransferase (GGT). Thus, better biomarkers of progressive ALD are needed. Furthermore, there is a great need for detailed information on the driving factors of the individual differences in the progression of ALD, which has been linked to multiple metabolic processes, including dyslipidemia, altered bile acids, increased gut permeability and dysbiosis of gut microbiota (Mathur et al. 2021, He et al. 2021, Dubinkina et al. 2017, Maccioni et al. 2020). Because of the multitude of potentially relevant pathways, a global (non-targeted) metabolomics analysis of the circulating metabolome could help to discover metabolic processes related to ALD progression.

Heavy alcohol use has been shown to alter circulating metabolome and gut permeability as well as diminish the ability of vital nutrients pass through the gut-blood barrier (de la Monte and J Kril 2014, Kärkkäinen et al. 2020, Jaremek et al. 2013, Lehikoinen et al. 2018, Würtz et al. 2016, Leclercq and Timary 2024, Kärkkäinen et al. 2024). Previous studies have shown that changes in the circulating metabolome are associated with liver cirrhosis in ALD patients (Bajaj et al. 2017, Xu et al. 2023) and metabolite profiles could be used to predict the development of various alcohol-associated diseases including ALD (Barrett et al. 2023, Kärkkäinen et al. 2020). However, the differences in the circulating metabolome between non-progressive and progressive ALD have not been extensively studied. Because progressive ALD has a significantly worse prognosis and higher mortality rate than non-progressive ALD, understanding these differences is important.

Our aim was to measure the circulating metabolite profiles associated with progressive ALD when compared to non-progressive ALD or healthy controls. We used serum samples collected from patients at the start of alcohol detoxification treatment and analyzed metabolite profiles using a non-targeted liquid chromatography mass spectrometry (LC-MS) based metabolomics method combined with a targeted measurement of short chain fatty acids (SCFAs).

## Materials and Methods

### Patients

The study cohort consisted of alcohol use disorder (AUD) patients who were admitted for standardized and controlled 3-week detoxification and rehabilitation program in Cliniques Universitaires Saint-Luc, Brussels, Belgium between 2017 - 2019. A healthy volunteer control group (n=32), who consume socially low amounts of alcohol (<20 g/day), was recruited separately. The patients had a longstanding history of alcohol abuse. Daily alcohol use was inquired from all the patients. They were heavy drinkers who consumed more than 60 g of alcohol per day and were actively drinking until the day of admission. Exclusion criteria included antibiotic use during the two months preceding enrollment, immunosuppressive medication, diabetes, BMI > 30, inflammatory bowel disease, known liver disease of any etiology other than ALD, and clinically significant cardiovascular, pulmonary or renal co-morbidities.

The participants were divided into groups depending on their clinical parameters (Table 2): a non-progressive ALD group (n=50) with controlled attenuation parameter (CAP) results > 250 dB/m, but normal liver enzymes (AST < 40 IU/l, ALT < 40 IU/l) and elastography measured liver stiffness results < 7.6 kPa, and a progressive ALD group (n=50) with CAP > 250 dB/m, increased AST and ALT levels and liver stiffness > 7.6 kPa. The non-progressive ALD group included patients with isolated steatosis. The progressive ALD group included patients with possible steatohepatitis or fibrosis.

On the day of hospital admission fasting serum and plasma EDTA samples were collected from the patients and Fibroscan^®^ (Echosense, Paris, France) measurements were performed. (Table 2). The sample collection protocol has been previously been described in detail (Maccioni et al. 2020). Standard biochemical analyses were obtained from the biochemistry laboratory of the hospital. Blood soluble CD14 (sCD14), M65 cell death biomarker (M65) and peptidoglycan-recognition proteins (PGRPs) were determined using commercially available kits (ELISA kits, Thermo Fisher Scientific, WA, USA,) (Table 2).

### Ethical considerations

The study complies with the Declaration of Helsinki and the Declaration of Istanbul. Written informed consent was obtained from all participants. The study has been approved by the “Comité d’éthique Hospitalo-facultaire Saint-Luc UCLouvain” (B403201422657). The reporting of this study has been done in accordance with the STROBE criteria.

### Non-targeted metabolomics analysis

The serum samples were sent to the University of Eastern Finland, Kuopio, Finland for non-targeted metabolomics analysis. The samples were stored at -80°C until use. When taken out of storage, they were thawed in ice water, after which they were kept in wet ice until assayed. The samples were then vortexed at the maximum speed with Vortez Genie 2 (Scientific industries, Bohemia, NY, USA). 400 μL of frigid acetonitrile was added to a 96-well plate with a filter plate. The vortexed serum samples were then added to the 96-well plate. Pooled quality control samples were done by collecting 10 μL of each sample and adding them to the same tube and mixing. By pipetting 4 times, the acetonitrile and samples were mixed. After all the samples were ready, the 96-well plate was centrifuged 700x g for 5 minutes at 4 °C with Heraus Megafuge 40R (Thermo Fisher Scientific). After the centrifuging, the filter plate was removed, and the plate sealed with the 96-well cap mat. Samples were prepared separately for reverse phase (RP) and hydrophilic interactions liquid chromatography (HILIC) analyses.

The serum samples were analyzed with non-targeted liquid chromatography mass spectrometry metabolomics using ultra-high performance liquid chromatography (UPLC) combined with Thermo Q Exactive^TM^ Hybrid Quadrupole-Orbitrap mass spectrometer (Thermo Scientific). Used reversed phase (RP) column was Zorbax Eclipse XDB-C18, particle size 1.8 µm, 2.1 × 100 mm (Agilent Technologies) and the used hydrophilic interaction chromatography (HILIC) column was Acquity UPLC BEH Amide 1.7 µm, 2.1 × 100 mm (Waters Corporation, Milford, Massachusetts, USA). The column temperature was 40 °C and the flow rate was 0.4 mL/min (mobile phase A: H2O + 0.1 % HCOOH, B: MeOH + 0,1 % HCOOH, 16.5 min gradient). for the RP mode and for the HILIC mode, the column temperature was 45 °C and the mobile phase flow rate was 0,6 mL/min (mobile phase A: 50% acetonitrile + 20 mM ammoniumformate buffer, B: 90 % acetonitrile + 20 mM ammoniumformate buffer, 12.5 min gradient). Positive and negative electrospray ionization (ESI) were used for HILIC and RP analytic modes. ESI ray voltage was 3.5 kV for positive and 3.0 kV for negative mode.

Metabolite identification, peak picking and alignment were done with MS-DIAL version 4.80 (Tsugawa et al. 2015). Preprocessing, including drift correction and missing value imputation, was done with ‘notame’ R-package (Klåvus et al. 2020). Metabolite identifications were ranked according to the community guidelines (Sumner et al. 2007). Metabolites in level 1 were matched against accurate mass, isotopic pattern, retention time, and product ion spectra (MSMS) of fragmented ions from the in-house library of chemical standards built using the same experimental conditions. Usage of the Level 2 includes metabolites with matching exact mass and MSMS spectra from public libraries, published papers or in the case of lipids, the built-in MS-DIAL library. Level 3 identification includes metabolites, whose chemical group has been recognized. All things equal, from multiple different ion forms of a certain compound, the most common product ion form was presented.

### Serum short chain fatty acids analysis

The analysis of serum acetic acid, propionic acid and butyric acid levels was based on previously published method (Nylund et al. 2020) with modifications. Serum samples stored at -80°C were thawed on wet ice and vortexed before processing in five batches. 150 µl of serum was aliquoted in a 10 mL vial holding a solution consisting of 0.5 g of NaH_2_PO_4_ and 1350 µL of cold Milli-Q (MQ) water. Analytical blank samples holding only 0.5 g of NaH_2_PO_4_ and 1500 µL of cold MQ water were prepared in an equivalent manner as study samples. Individual stock solutions (500–2500 ppm) of acetic, propionic and butyric acids (Sigma-Aldrich, Saint Louis, Missouri, USA) prepared by dissolving standards in MQ water. Pooled analytical standard was prepared by combining 25 µL of each stock solution in a 10 mL vial holding a solution consisting of 0.5 g of NaH_2_PO_4_ and 1500 µL of cold MQ water. Vials were gently whirled and placed on the autosampler until analysis. Three injections consisting of blank, analytical standard and blank were injected in the beginning, end and every 26 injections during the sequence.

Solid-phase microextraction coupled to gas chromatography and mass spectrometry (SPME-GC-MS) analysis was carried out on a Thermo Trace 1310 – TSQ 8000 Evo instrument holding an TriPlus RSH autosampler (Thermo Scientific) kept at + 4°C throughout the analysis. SCFAs were extracted using a 75 µm CAR/PDMS Fused Silica SPME fiber (Supelco, Bellefonte, PA, USA) that was conditioned according to the manual. Samples were incubated for 10 minutes followed by an extraction time of 40 minutes. Incubation and extraction temperatures were kept at 40°C. Five-minute desorption temperature was 240 °C in the GC injector port with a splitless mode. Chromatographic separation was performed by fused silica capillary column (Supelco) SPB-624 (60 m × 0.25 mm × 1.4 µm) under a carrier gas (helium) 1.40 mL/min. The total GC oven program time was 48 min where initial temperature was held at 40°C for 10 minutes, ramped by 5°C/min to 200 °C and then held at 200 °C for 10 min. MS was operated at 240 °C and electron ionization voltage of 70 eV and ions were scanned in the full scan mode (30–300 amu). The instrument was operated, and data was analyzed with Chromeleon 7.2.10 software (Thermo Fisher Scientic) by comparison of retention times and peak intensiMSties against SCFA external analytical standards. Manually integrated spectral areas were exported to spreadsheet format for statistical analysis.

### Statistical analysis

For the statistical analysis, the results from the metabolomics and the SCFA analyses were combined. Welch’s one-way analysis of variance (ANOVA) was used to determine which metabolites had significant differences between the three study groups. For group-to-group comparison, Welch’s t-test and Cohen’s d effect sizes were used. For the group comparisons, the α level was adjusted to 0.0005 to account for multiple testing, based on the number of latent components (106) needed to explain 95% of variance in the metabolomics data in a principal component analysis (PCA). Multivariate analysis of the differential molecular features, partial least sum of squares discriminant analysis (PLS-DA) was used and variable importance for projection (VIP) values are reported. For correlation, the Spearman method was used with false discovery rate (FDR, cutoff 5%) to account for multiple testing. Correlation analysis was done for all identified metabolites of identification level of 1 or 2. The data was analyzed with R (version 4.2), R-studio (version 492), JASP (version 0.16.2.0) (University of Amsterdam, Netherlands) and Microsoft Excel (Microsoft, Redmond, Washington, USA). The pivotal packets in use were: notame, missforest, lme4 and Imertest. Graphs were drawn with JASP (version 0.16.2.0) and GraphPad Prism (version 9) (Graphpad Software, La Jolla, California, USA).

## Results

### Study population

The demographic, biochemical and clinical data are shown in Tables 1 and 2, respectively. The two ALD groups were similar in terms of age, sex and BMI (Table 1). However, the BMI and age were significantly lower in the control group compared to the ALD groups. According to the selection criteria, liver stiffness, AST- and ALT-levels were significantly different between the two ALD groups (Table 2). The INR, bilirubin, and albumin values were within the normal range in the two ALD groups.

**Table 1.** Demographical characteristics of the study populations.

|  | **Progressive**<br>**ALD (n=50)** | **Non-**<br>**progressive**<br>**ALD (n=50)** | **Healthy**<br>**controls (n=32)** | **p-value** |
| --- | --- | --- | --- | --- |
| **Sex** (n%) | 16 women (32%) | 17 women (34%) | 16 women (50%) | 0.218<sup>a</sup> |
| **Age** (years,<br>Mean ± SD) | 51.1 ± 9.9 | 48.3 ± 11.3 | 40.6 ± 14.6 | 0.001<sup>b</sup> |
| **BMI** (kg/m<sup>2</sup>,<br>Mean ± SD) | 25.8 ± 3.9 | 24.3 ± 3.4 | 23.7 ± 3.7 | 0.036<sup>b</sup> |

Legend: ALD = alcohol-related liver disease, BMI = body mass index, <sup>a</sup> Chi<sup>2</sup> test, <sup>b</sup> Welch's ANOVA.

**Table 2.** Baseline biochemical, fibroscan and bacterial translocation markers measurements and inquired daily alcohol intake from the alcohol-related liver disease (ALD) groups, mean and standard deviation shown.

|  | **Progressive**<br>**ALD (n=50)** | **Non-**<br>**progressive**<br>**ALD (n=50)** | **Welch's t-test** | **Cohen's effect**<br>**size** |
| --- | --- | --- | --- | --- |
| **AST** (IU/L) | 119.5 ± 85.3 | 26 ± 7.5 | p<0.001 | d=1.54 |
| **ALT** (IU/L) | 87.2 ± 62.1 | 23.55 ± 9.82 | p<0.001 | d=1.43 |
| **AST/ALT-ratio** | 1.5 ± 0.71 | 1.2 ± 0.35 | p=0.004 | d=0.61 |
| **GGT** (IU/L) | 443.6 ± 477.7 | 58.10 ± 43.8 | p<0.001 | d=1.18 |
| **M65** (IU/L) | 687.5 ± 534.6 | 155.7 ± 79.1 | p<0.001 | d=1.50 |
| **Bilirubin** | 0.78 ± 0.84 | 0.52 ± 0.26 | p=0.041 | d=0.42 |
| **Albumin** (gr/L) | 46.1 ± 6.6 | 47.3 ± 4.1 | p=0.290 | d=0.22 |
| **INR** | 1.1 ± 0.2 | 1.0 ± 0.1 | p=0.020 | d=0.50 |
| **CAP** (dB/m) | 308.5 ± 48.6 | 255.3 ± 56.9 | p<0.001 | d=1.01 |
| **Liver Stiffness**<br>(kPa) | 15.9 ± 15.4 | 4.4 ± 1.1 | p<0.001 | d=1.05 |
| **sCD14** (ng/ml) | 2104.7 ± 446.8 | 1860.4 ± 318.5 | p=0.015 | d=0.59 |
| **PGRPs** (ng/ml) | 38.4 ± 19.6 | 43.0 ± 24.7 | p=0.387 | d=0.21 |
| **Alcohol intake**<br>(gr/day) | 208.3 ± 111.1 | 174.7 ± 105.1 | p=0.125 | d=0.311 |

Legend: ALT = alanine aminotransferase, AST = aspartate aminotransferase, GGT = gamma-glutamyl aminotransferase, CAP = controlled attenuation parameter, INR = International normalized ratio. PGRPs = peptidoglycan recognition proteins, M65 = cell death biomarker M65, sCD14 = soluble CD14.

### Comparisons between ALD groups and controls

In the non-targeted metabolomics analysis, we collected 5253 molecular features with the two RP modes and 1824 molecular features with the two HILIC modes (Supplementary Table 1,). Levels of acetic acid, propionic acid and butyric acid were detected with the targeted SCFA-analysis. In the ANOVA comparison, 3461 molecular features had a p-value <0.05. Furthermore, 1868 molecular features had p-value under the corrected α-level accounting for multiple testing (p<0.0005). Excluding multiple molecular features for a taken compound, 111 identified metabolites had a p-value under the multiple testing corrected α-level.

The total 111 identified metabolites and 3 short chain fatty acids had significant differences between the control group and the ALD groups (Figures 1 and 2). Glutamic acid, hypoxanthine, lysophosphatidylcholines (LPC), phosphatidylcholines (PC), fatty acids (FA) and acylcarnitines (CAR) all had significant differences between the control group and the two ALD groups (Figures 1 and 2). Furthermore, sphingomyelin (SM) levels were lower in both the non-progressive ALD and progressive ALD groups when compared to the control group (Figure 2). Lysophosphatidylethanolamine (LPE) levels were higher in the ALD groups when compared to the control group. 5-aminovaleric acid betaine (5-AVAB) was significantly increased in the progressive ALD group in comparison to the control group (p=0.0002, d=0.56), but did not reach significance when comparing the non-progressive ALD group to the control group or in the ALD groups intergroup comparison.

**Figure 1.**
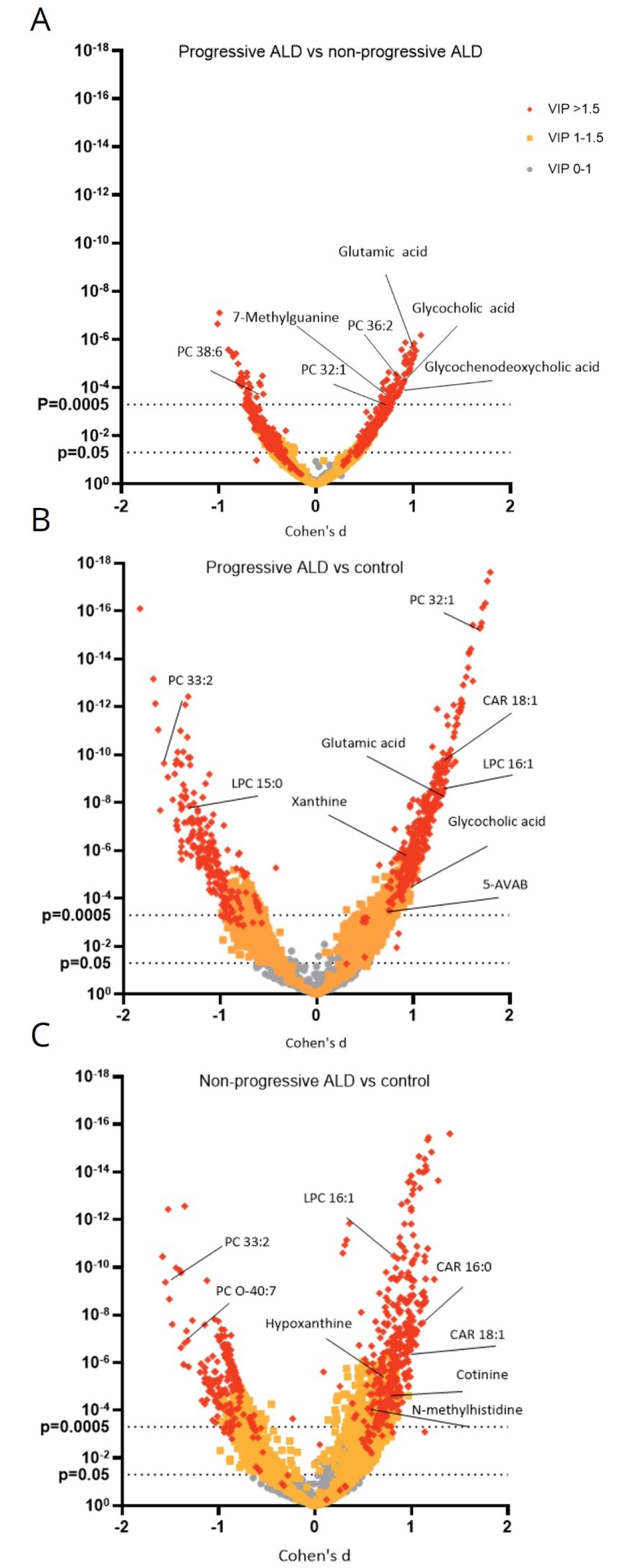
Volcano plot of all molecular features stratified by their significance in group comparison of the three different groups. Cohen’s d effect sizes and p-values from Welch’s t-test are shown for comparison between progressive alcohol-related liver disease (ALD) and non-progressive ALD (A), progressive ALD and healthy controls (B) and non-progressive ALD and healthy controls (C). Control = control group, VIP = variable importance in projection, LPC = lysophosphatidylcholine, PC = phosphatidylcholine, CAR = acylcarnitine.

**Figure 2.**
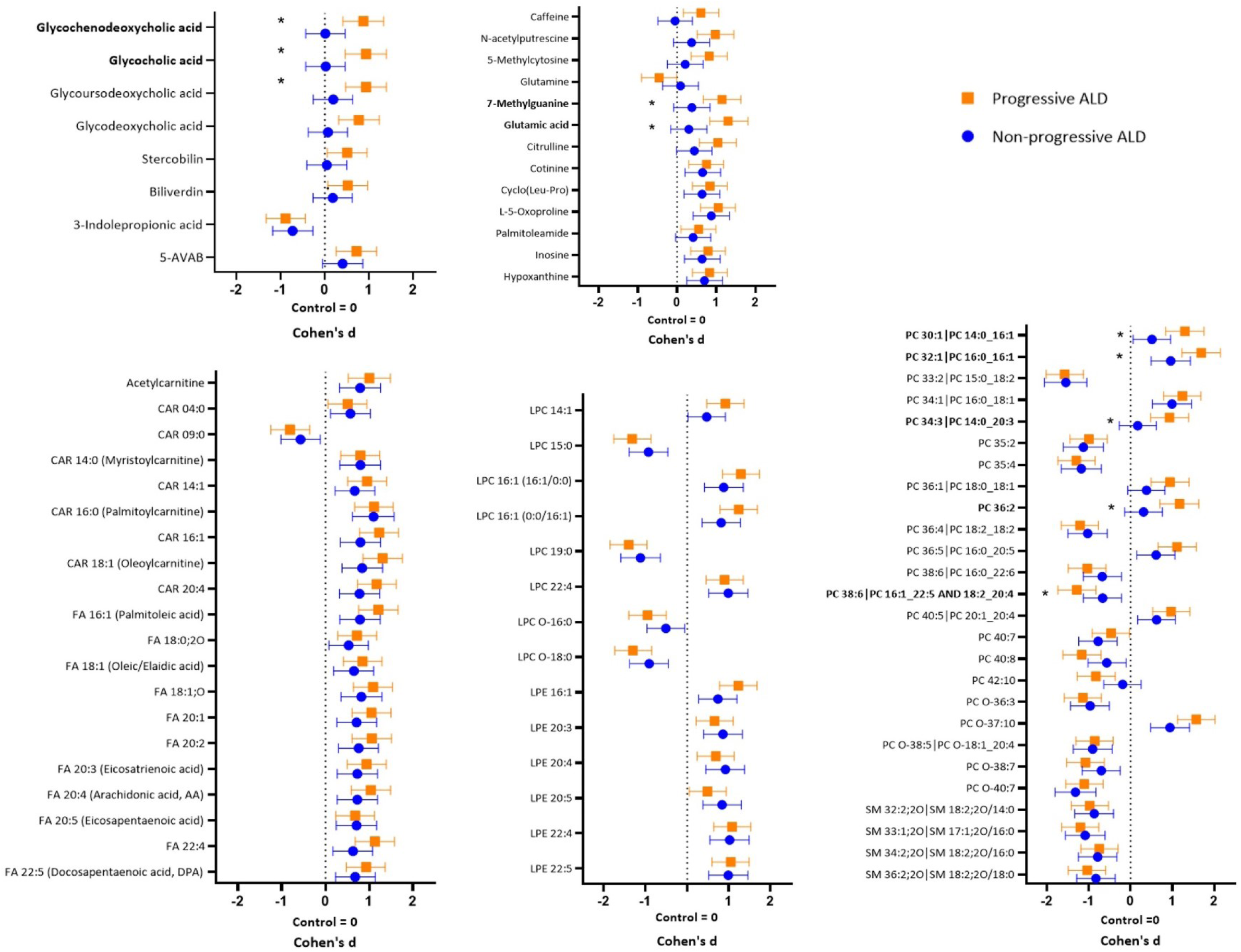
Changes in the significantly differential identified metabolites between alcohol-related liver disease (ALD) groups and metabolites of interest in comparison to healthy controls. Cohen’s d values with 95% confidence intervals are shown for comparisons between the control group and both progressive ALD and non-progressive ALD groups. Metabolites which have a significant difference (p<0.0005) between non-progressive ALD and progressive ALD group are marked with * and bolded. Control = control group, ALD = alcohol-related liver disease, FA = Fatty acid, SM = sphingomyelin, CAR = acylcarnitine, PC = phosphatidylcholine, LPC = lysophosphatidylcholine, LPE = lysophosphatidylethanolamine. 5-AVAB = 5-amino valeric acid betaine.

### Comparison between progressive and non-progressive ALD groups

When comparing the progressive ALD group to the non-progressive ALD group, the levels of glycocholic acid (p<0.0001, Cohen’s d=0.91), glycochenodeoxycholic acid (p<0.0001, d=0.90) (Figure 1 and 2), glutamic acid (p<0.0001, d=1.01), and 7-methylguanine (p=0.0002, d=0.77) (Figure 2) were increased. Furthermore, PC 14:0_16:1 (p=0.0001, d=0.80), PC 16:0_16:1 (p<0.0001, d=0.73), PC 14:0_20:3 (p=0.0002, d=0.76), and PC 36:2 levels (p<0.0001, d=0.85) were higher in the progressive ALD group when compared to the non-progressive ALD group (Figure 2). In contrast, PC 38:6 (16:1/22:5) (p=0.0002, d=-0.61) levels were lower in the progressive ALD group when compared to non-progressive ALD group (Figure 1 and 2).

### Correlations

Correlation analysis with background variables (Table 1 and Table 2) was done for all identified metabolites and short chain fatty acids (Supplementary Table 1). A total of 139 metabolites of identification levels 1 and 2 and short chain fatty acids had significant correlations (FDR corrected p-value < 0.05) with at least one background variable (Figure 3).

**Figure 3.**
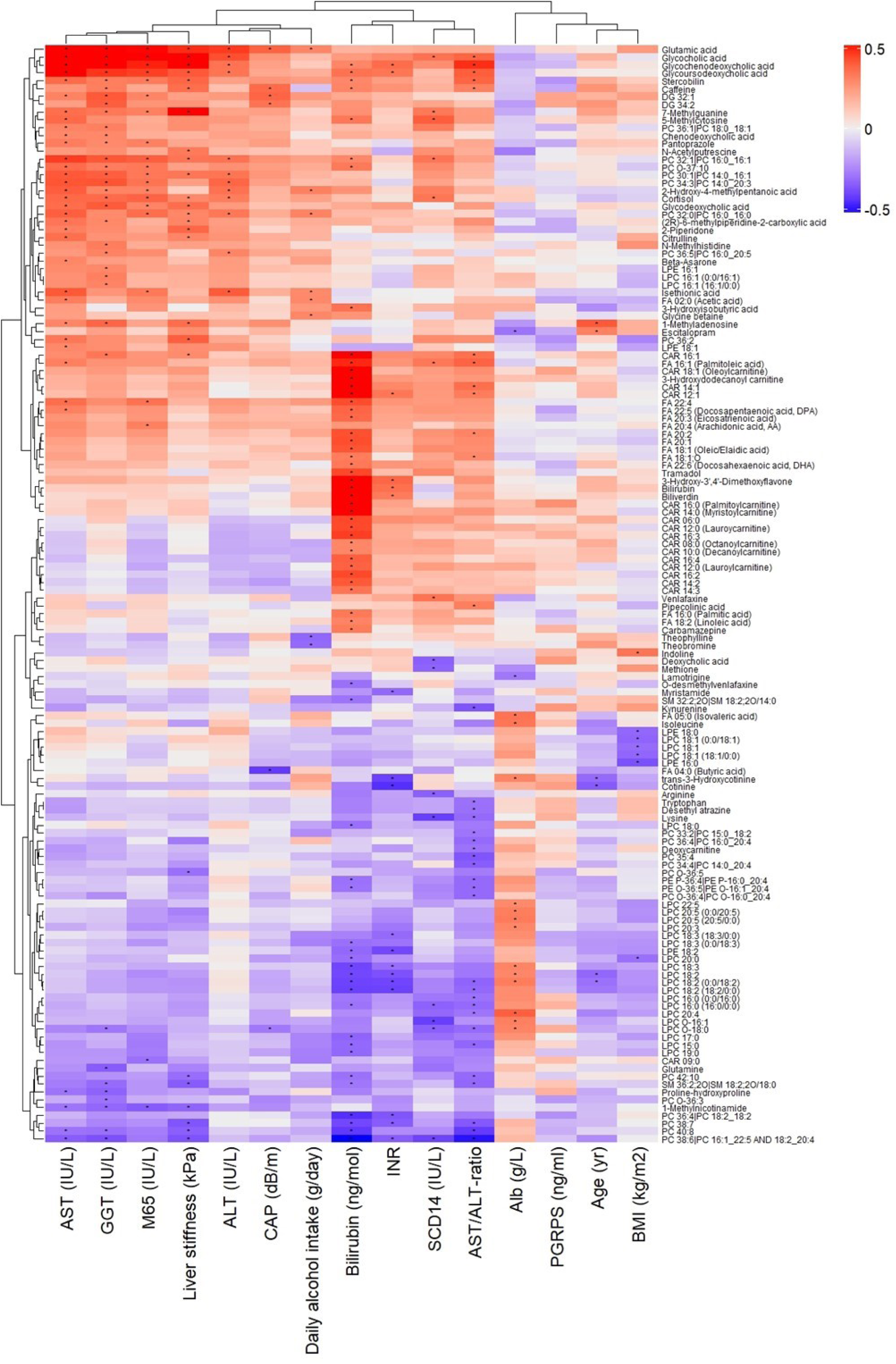
Spearman correlations between identified metabolites and clinical parameters with at least one significant correlation. The color scale slides with the correlation coefficient, ones with positive correlation become increasingly red according to the correlation coefficient and those that have a negative correlation coefficient become increasingly blue. Alb = albumin, ALT = alanine aminotransferase, AST = aspartate aminotransferase, BMI = body mass index, CAP = controlled attenuation parameter, GGT = gamma glutamyltransferase, INR = international normalized ratio, PGRPS = peptidoglycan recognition proteins, sCD14 = blood soluble CD14. CAR = acylcarnitine, DG = diglyceride, FA = Fatty acid, SM = sphingomyelin, LPC = lysophosphatidylcholines, LPE = lysophosphatidylethanolamine, PC = phosphatidylcholine, PE = phosphatidylethanolamine. * FDR corrected p-value < 0.05.

Glycine conjugated bile acids, multiple phospholipids, 7-methylguanine, 5-methylcytosine and stercobilin correlated with AST. Similarly, ALT had a positive correlation with glycine conjugated bile acids and phospholipids, but not with 7-methylguanine, 5-methylcytosine and stercobilin. Accordingly, the AST/ALT-ratio correlated positively with glycine conjugated bile acid levels and negatively with PCs and LPCs. GGT correlated positively with glutamic acid, glycine conjugated bile acids, PCs, LPCs and negatively with glutamine. Alcohol use had negative correlations with theophylline and theobromine and positive correlations with PC 32:0|PC 16:0_16:0, glutamic acid, glycine betaine, acetic acid (FA 02:0), isethionic acid, and 2-hydroxy-4-methylpentanoic acid.

The international normalized ratio (INR) had significant negative correlations with multiple LPCs and positive correlations with biliverdin, bilirubin, CAR 12:1, glycoursedeoxycholic acid and glycochenodeoxycholic acid. Blood albumin had positive correlations with LPCs, isoleucine, isovaleric acid, trans-3-hydroxycotine and negative correlations with lamotrigine and escitalopram. Bilirubin correlated positively with CARs, FAs, biliverdin, stercobilin, glycochenodeoxycholic acid, glycoursedeoxycholic acid, 5-methylcytosine, caffeine, tramadol, 3-Hydroxyisobutyric acid and 3-hydroxy-3’ 4’-dimethoxyflavone. Bilirubin had negative correlations with multiple LPCs, SMs, PCs, O-desmethylvenlafaxine and LPE 18:2. sCD14 had a negative correlation with LPC O-16:1, LPC O-16:0, LPC O-18:0, and PC 38:6. Glycocholic acid, 7-mehtylguanine, 5-methylcytosine and PC 32:1 had positive correlations with sCD14. The PGRPs did not have significant correlations with any identified metabolites.

The only metabolites which reached significance in correlation analysis with the CAP values were positive associations with glutamic acid, caffeine, DG 32:1, DG 34:2 and a negative correlation with butyric acid and LPC 0-18:0 (Figure 3). Elastography liver stiffness results correlated positively with glycine bound bile acids, glutamic acid, stercobilin, caffeine and 7-methylguanine among others. Conversely, liver stiffness had a negative correlation with 1-methylnicotinamide and several PCs. BMI correlated positively with indoline and negatively with LPE 16:0, LPE 18:0, LPC 18:1 (0:0/18:1), LPC 18:1 (18:1/0:0), LPC 20:0. Age correlated positively with escitalopram and 1-methyladenosine levels and negatively with trans-3-hydroxycotinine, LPC 18:2 (18:2/0) and LPC 18:2 (0:0/18:2).

## Discussion

The results of this study demonstrated distinct differences in the serum metabolite profiles between patients with progressive ALD and those with non-progressive ALD. Increased levels of glycine conjugated bile acids, glutamate, PC 14:0_16:1, PC 16:0_16:1, PC 14:0_20:3, PC 36:2 and 7-methylguanine demonstrated metabolic and enterohepatic alterations already early in the ALD disease progression, before the patients develop liver decompensation based on clinical and biochemical parameters (normal INR, albumin and bilirubin levels).

The primary finding was that glycine conjugated bile acids were significantly increased only in the progressive ALD group, when compared to the non-progressive ALD group and controls. Additionally, glycine conjugated bile acid levels also correlated strongly with liver stiffness measured with elastography. Increased levels of glycine conjugated bile acids have been reported previously in severe forms of ALD such as severe alcohol-associated hepatitis and cirrhosis (Brandl et al. 2018, Ciocan et al. 2018). Our results indicate that these bile acids may already contribute to disease progression at much earlier stages and therefore represent a potential target for therapy. However, most bile acids in serum are conjugated with either glycine or taurine (Hofmann 1999) and the conjugated bile acids are not directly hepatotoxic whereas non-conjugated bile acids are (Kundu et al. 2015, Woolbright et al. 2015). Therefore, the potential role of glycine conjugated bile acids as drivers or accelerators of disease progression is more complex. Primary bile acids cholic acid and chenodeoxycholic acid increased in the progressive ALD are produced by the neutral pathway initiated by 7α-hydroxylase (CYP7A1) in the liver (Farooqui et al. 2022). Secondary bile acids, resulting from transformation of primary bile acids by intestinal bacteria, were also increased, (ursodeoxycholic acid, Chen et al. 2021). This indicates that both the internal production of primary bile acids, as well as the production of secondary bile acids by the gut microbiota is altered in progressive ALD. Alcohol use has been shown to increase bile acid levels in the liver of early-stage ALD mice model (Charkoftaki et al. 2022) as well as bile acid formation in hepatocytes *in vitro* (Nilsson et al. 2007). Heavy alcohol use has previously been shown to increase levels of primary glycine-conjugated bile acids in plasma (Muthiah et al. 2022). Alcohol use can also alter gut microbiota further affecting the enterohepatic circulation of bile acids (Liu et al. 2022). Bile acids have also been linked to gut permeability, intestinal immunity, glutamine and glutamic acid metabolism, lipid metabolism and more broadly to energy metabolism through the activation of farnesoid X -receptor (FXR) and secondary pathways (Chiang and Ferrel 2022, Keitel et al. 2010, Renga et al. 2011). It is conceivable that the high glycine conjugated bile acid levels could also be linked to the increased glutamic acid and altered phospholipid levels seen in the progressive ALD group.

7-methylguanine was significantly higher in the progressive ALD group than in the other two groups, possibly in reaction to liver inflammation. It is a nucleic acid metabolite that inhibits poly(ADP-ribose)polymerase-1 and poly(ADP-ribose)polymerase 2. Increased serum levels of 7-methylguanine have been previously linked to increased risk of hepatocellular carcinoma (Stepien et al. 2021), but it’s role in the progression of ALD is unclear.

In our study glutamic acid levels were positively correlated with alcohol consumption, as well as with ALD progression. Previous studies have shown that alcohol use in general increases circulating glutamic acid levels (Holmes et al. 2013, Lehikoinen et al. 2018, Heikkinen et al. 2019). Glutamic acid is a part of multiple energy metabolism pathways and functions as an excitatory neurotransmitter (Brosnan and Brosnan 2013). Glutamic acid has also been linked to alcohol hepatotoxicity as an enhancer of mitochondrial oxidative stress (Teplova et al. 2017) and therefore high glutamic acid levels could accelerate the progression of liver damage.

The major differences in the serum levels of FAs, CARs, PCs, and LPCs observed between the ALD groups and the control group are in line with earlier findings of alterations in lipid profiles in a variety of profiles in steatotic liver diseases (Paul et al. 2022, Enooku et al. 2019, Raja et al. 2021). Sphingomyelin levels in the ALD patients were lower than those in the control group, but there was not significant difference between the two ALD groups. Sphingomyelin depletion has been linked to the progression of liver-related events and ALD without metabolic syndrome overlap (Thiele et al. 2023). Overall, our results confirm this association since patients with diabetes and BMI>30 were excluded from the study leaving a study population of primarily ALD patients with fewer metabolic risk factors. Of the SMs, only SM 36:2 had a significant correlation with liver stiffness results, indicating that sphingomyelin depletion might be more characteristic of inflammation than fibrosis. Furthermore, both phosphatidylcholines and lysophosphatidylcholines showed significant differences between the study groups. Phosphatidylcholines have been proposed to play a role in the progression and pathogenesis of metabolic dysfunction-associated steatotic liver disease (MASLD, previously NAFLD) (Sherriff et al. 2016). Our results raise the possibility that this might also be the case in the earlier stages of ALD in a population without features of metabolic syndrome. Alcohol intoxication in patients with ALD has been shown to decrease circulating free FAs and lysophosphatidylcholines and increase circulating triglyceride levels (Israelsen et al. 2021). In this study fatty acid levels were significantly elevated in the ALD groups in comparison to the healthy control group, suggesting that the baseline levels of FAs are higher in ALD when compared to people without liver disease.

Interestingly, high serum levels of the microbiota-associated metabolite 5-aminovaleric acid betaine (5-AVAB), which has previously been associated with obesity and MASLD in both humans and mouse models (Liu et al. 2021, Haikonen et al. 2022), were seen in the progressive ALD group when compared to the controls. This provides a potential link between the altered intestinal microbiota observed in the development and progression of AUD and ALD. *Bifidobacteria* and *coriobacteriaceae* have been linked to 5-AVAB levels (Haikonen et al. 2022). 5-AVAB influences lipid metabolism by reducing the β-oxidation of FAs via an inhibition of the cell membrane carnitine transporter (Kärkkäinen et al. 2018, Haikonen et al. 2022). Consequently, reduced β-oxidation could explain some of the changes observed in lipids.

This study had some limitations. The study was cross-sectional with a relatively small sample size. Therefore, to validate and increase the generalizability of the results, they should be validated in larger cohorts with a longitudinal design. In addition, the healthy control group was significantly younger than the two ALD groups, which could have an impact on the cross-group comparisons. Alcohol consumption was based on self-reported daily alcohol use. Likely, using the timeline follow-back method to assess ethanol consumption would have been more accurate. Furthermore, as in most non-targeted metabolomics studies, many significantly altered molecular features were not matched with known substances (Supplementary Table 1). In conclusion, patients with early-stage progressive ALD with no liver decompensation have altered metabolic and enterohepatic processes when compared to patients with non-progressive ALD. If the present findings of increased serum levels of glycine conjugated bile acids, and glutamic acid in progressive ALD can be validated in longitudinal prospective cohorts, these alterations could serve as potential early phase biomarkers and/or treatment targets for predicting and preventing the progression of ALD.

## Data Availability

The data that support the findings of this study are available on request from the corresponding author, OK.

## List of abbreviations

ALD: alcohol-related liver disease
ANOVA: analysis of variance
AUD: alcohol use disorder
BMI: body mass index
CAP: controlled attenuation parameter
CAR: acylcarnitine
CYP7A1: 7α-hydroxylase
DG: diglyceride
ESI: electrospray ionization
FA: Fatty acid
FDR: false discovery rate
FXR: farnesoid X-receptor
HILIC: hydrophilic interactions liquid chromatography
INR: international normalized ratio
LC-MS: liquid chromatography mass spectrometry
LPE: lysophosphatidylethanolamine
LPC: lysophosphatidylcholine
M65: M65 cell death biomarker
PC: phosphatidylcholine
PCA: principal component analysis
PE: phosphatidylethanolamine
PGRPs: peptidoglycan-recognition proteins
PLS-DA: partial least sum of squares discriminant analysis
RP: reversed phase
sCD14: blood soluble CD14
SPME-GC-MS: solid-phase microextraction coupled to gas chromatography and mass spectrometry
UPLC: ultra-high performance liquid chromatography
VIP: variable importance in projection
5-AVAB: 5-aminovaleric acid betaine

## Acknowledgements

We thank Miia Reponen and Juulia Kuparinen for technical assistance with the mass spectrometry analyses. The authors also thank Biocenter Finland and Biocenter Kuopio for supporting the UEF core LC-MS laboratory facility.

